# Kalman Filter Based Short Term Prediction Model for COVID-19 Spread

**DOI:** 10.1101/2020.05.30.20117416

**Authors:** Suraj Kumar, Koushlendra Kumar Singh, Prachi Dixit, Manish Kumar Bajpai

## Abstract

COVID-19 has emerged as global medical emergency in recentdecades. The spread scenario of this pandemic has shown many variations. Keeping all this in mind, this article is written after various studies and analysis on the latest data on COVID-19 spread, which also includes the demographic and environmental factors. After gathering data from various resources, all data are integrated and passed into different Machine Learning Models to check the fit. Ensemble Learning Technique,Random Forest, gives a good evaluation score on the test data. Through this technique, various important factors are recognised and their contribution to the spread is analysed. Also, linear relationship between various features is plotted through heatmap of Pearson Correlation matrix. Finally, Kalman Filter is used to estimate future spread of COVID19, which shows good result on test data. The inferences from Random Forest feature importance and Pearson Correlation gives many similarities and some dissimilarities, and these techniques successfully identify the different contributing factors. The Kalman Filter gives a satisfying result for short term estimation, but not so good performance for long term forecasting. Overall, the analysis, plots, inferences and forecast are satisfying and can help a lot in fighting the spread of the virus.

## 1. Introduction

Corona virus (COVID-19) belongs to a group of viruses that cause disease in mammals and birds. The name ‘corona virus’ is derived from Latin word corona meaning “crown”. The name refers to the characteristic appearance of virions by electron microscopy. This is due to the ‘viral’ spikes which are proteins on the surface of the virus. It causes respiratory throat infections in humans. COVID-19 is large spherical particle with surface projections. The average diameter is approximately 120 nanometer (nm) [1]. Envelope diameter is 80 nm while spikes are 20 nm long. The viral envelope consists of a lipid out layer where the membrane, envelope and spike structural proteins are anchored. Inside the envelope, there is the nucleocapsid, which is formed of proteins, single stranded RNA genome. COVID-19 contains SSRNA [2]. The genome size ranges from 26.4–31.7 ab. It also contains “polybasic cleavage site” which increases pathogenicity and transmissibility. The spike protein is responsible for allowing the virus to attach and fuse with the membrane of a host cell. COVID-19 has sufficient affinity to the receptor angiotensin converting enzyme on human cells to use them as mechanism of cell entry [3]. The cell’s trans-membrane protease serine cuts open the protein and exposing peptide after a COVID-19 virion attaches to a target cell. The virion then releases RNA into the cell and forces the cell to produce and disseminate copies of the virus [4]. The RNA genome then attaches to host cells ribosome for translation. The basic reproductive number (RO) of the virus has been estimated to be between 1.4 to 3.9. The infection transmits mainly through droplets released from infected person or surfaces containing these infected droplets [5].

The detection and controlling of any infection based disease outbreaks have been major concern in public health [2]. The data collection from different sources such as health departments, emergency department, weather information etc plays vital role in decision making to control the epidemic. It has been well established in literature that data sources contains important information that helps to current public health status [6, 7]. Now, it becomes important that world health authorities and the global population remain vigilant against a resurgence of virus. Situational Information in social media during COVID-19 has been studied [8]. Markov switching model has been used to detect the disease outbreak [9]. Prediction model of HIV incidence has been proposed using neural network [10]. There are many methods exiting in literature to early detection of different diseases [11, 12, 13].

In case of any epidemic, the early prediction play vital role to control the epidemic. The Government agencies as well as public health organizations will prepare early according to the prediction. There are many prediction algorithms available in the literature for different types of data [2, 9]. Kalman filter is one of the popular filter to study of multivariable systems, highly fluctuated data, time varying systems and also suitable to forecast random processes [14]. Yang and Zhang used Kalman filter for prediction of stock price [15]. They have used Changbasihan as a test case to predict the stock price [15]. The extended Kalman filter in nonlinear domain has been studied by Iqbal et al [16]. Kalman filter is very useful in the field of Robotics [19]. It is extensively used to optimize the robotic movements, tracking of robots and their localization [19]. Kalman filter is also used in supply chain as abstraction [17]. It is also useful in manufacturing process to improve the capability of overall process [18]. Another useful application of Kalman filter was reported in literature to estimate parameters of train that is coating on a flat track [19].

The structural and replication mechanism followed by COVID-19 have motivated the study of demogramic and environmental factors affecting its spread. The present manuscript has included many such factors for the study e.g. Minimum Temperature, Maximum Temperature, Humidity, Rain Fall etc. The effect of these factors has been studied thoroughly on the spread rate. The effect is also studied on death rate and active cases rate.

## 2. Proposed Methodology

The Kalman filter tries to predict the state*X* ∈ *R^n^* of a discrete-time dependent process that is controlled by linear stochastic difference equation:

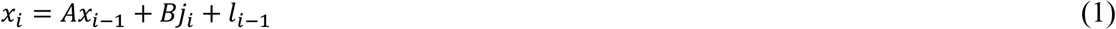

With a measurement*Y* ∈ *R^m^* which is

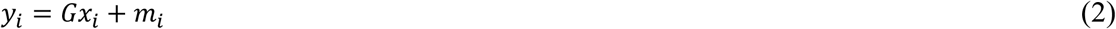

The random variable *l_i_* represents the process noise and *m_i_* represents the measurement noise. These both variables are assumed to be independent of each other with normal probablity distributions

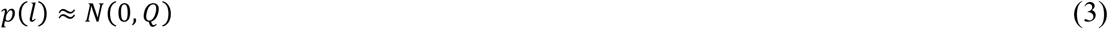

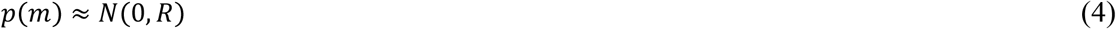

Here, *Q* is the process noise covariance matrix and *R* is the measurement noise covariance matrix. *A* is *nxn* matrix which establishes relationship between the state at the previous time step and the state at the current time step, when the driving function or process noise is absent. *B* isn*x*1matrix which establishes relationship between the optional control input *j* ∈ *R*^1^ and the state *x. G* is *mxn* matrix which establishes relationship between the state and the measurement*y_i_*.

### The Discrete Kalman Filter Algorithm

We can narrow the focus to the specific equations and their use in this version of the filter. The Kalman filter does estimation of a process by a kind of feedback control: the filter predicts the process state at some time and then accepts feedback in the form of measurements. Hence, the Kalman filter algorithm can be divided into two parts: *(i)* Time Update Equations (*Prediction Phase*) and *(ii)* Measurements Update Equations (*Feedback Phase*).

The time update equations are accountable for advancing the current state, and error covariance forward in time, to obtain a priori estimates of the upcoming time step. The measurement update equations are accountable for the feedback, i.e., fetching the actual measurement and changing the parameters to improve the Kalman Filter, in order to improve the posteriori estimates. Hence, in simple words, Time Update Equations can be considered as predictor equations, while the Measurement Update Equations can be considered as corrector equations, and the algorithm works in these two steps.

*Time Update Equations:*

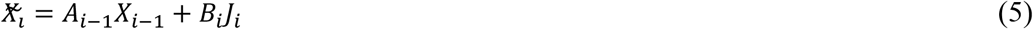

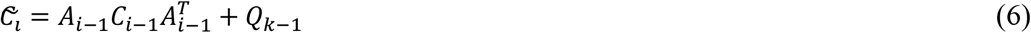

*Measurements Update Equations:*

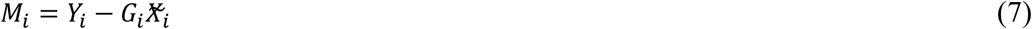

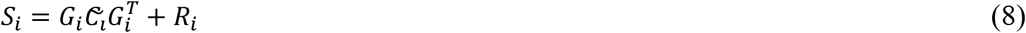

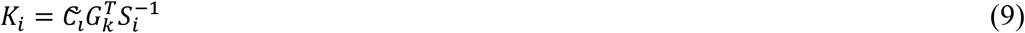

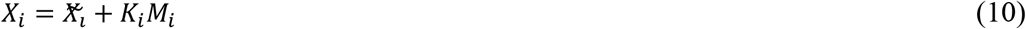

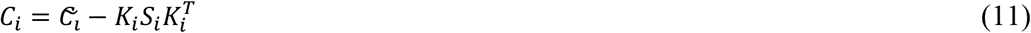

Here, 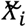 is the predicted mean on time step *i* before seeing the measurement and X_i_ is the estimated mean on time step *i* after seeing the measurement. 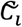 is the predicted covariance on time step *i* before seeing the measurement and *C_i_* is the estimated covariance on time step *i* after seeing the measurement. *Y_i_* is mean of the measurement on time step *i*. *M_i_* is the measurement residual on time step *i*. *S_i_* is the measurement prediction covariance on time step *i*. *K_i_* reflects how much prediction needs correction on time step *i*.

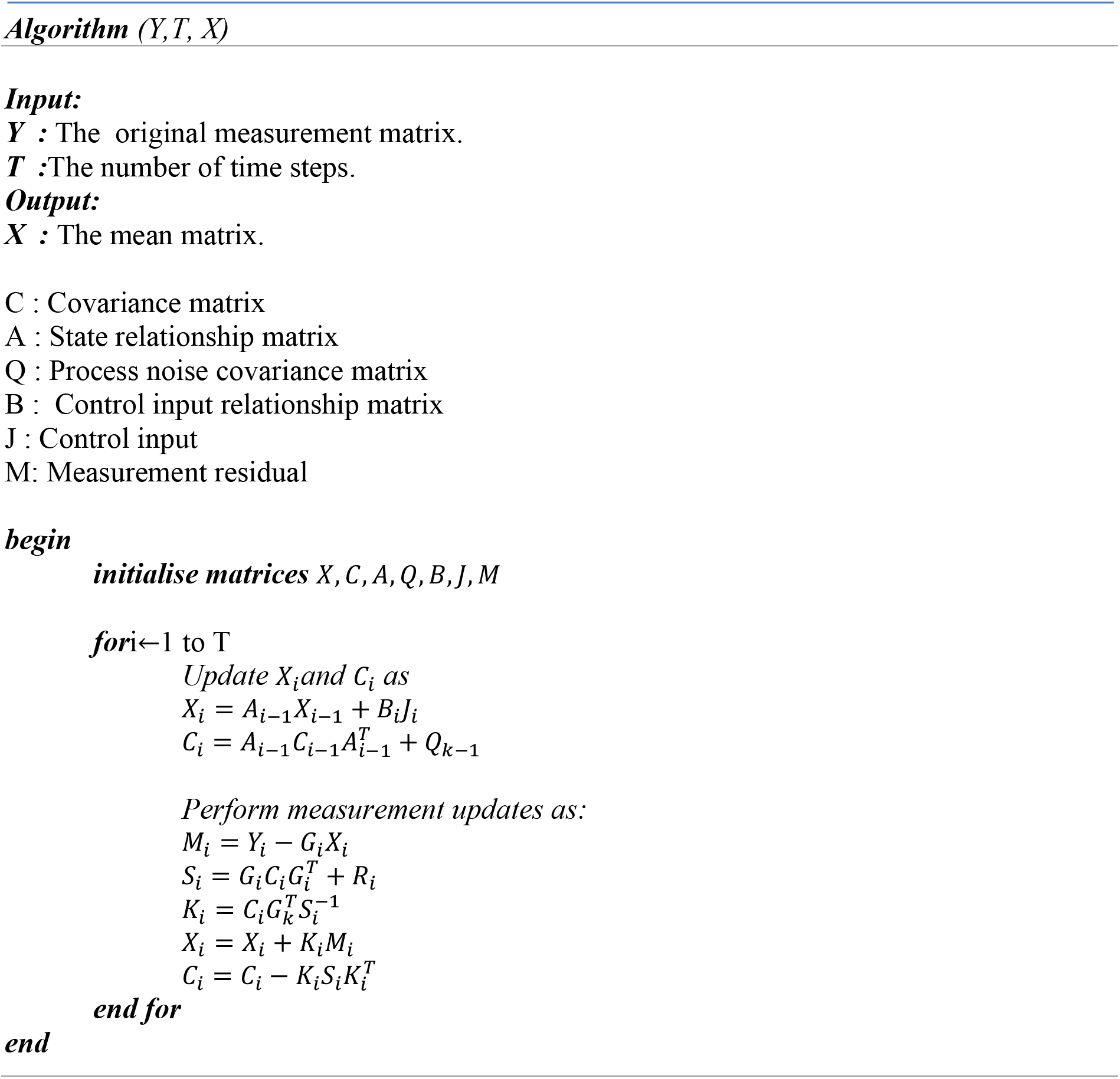

### Pearson Correlation

Correlation is a statistical technique that can show whether and how strongly pairs of variables are related. We have used Pearson Correlation Coefficient for analysing the relationship between the generated features and the number of confirmed cases. Pearson correlation coefficient is a statistic which lies between −1 to +1, and measures linear correlation between any two variables *Z* and *V*. +1 means total positive linear correlation, −1 mean total negative linear correlation and 0 means no correlation.

Pearson’s correlation coefficient can be presented as ratio of the covariance of the two variables Z and V and the product of their standard deviations.

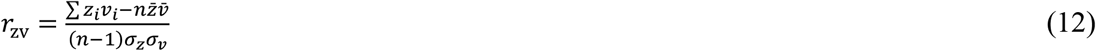

Here, *n* is sample size. *z_i_* mean. *v_z_* and *v_i_* and *v_i_* are individual sample points indexed with *i*. 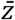 and 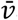 are sample mean. *σ_z_* and *σ_v_* are sample standard deviation.

### Feature Importance through Ensemble Learning Models

Decision Trees are an important type of algorithm for predictive modelling machine learning. The classical decision tree algorithms have been around for decades and modern variations like random forest are among the most powerful techniques available. A decision tree is created by dividing up the input space. A greedy approach is used to divide the space called recursive binary splitting. This is a numerical procedure where all the values are lined up and different split points are tried and tested using a cost function. The split with the best cost is selected. All input variables and all possible split points are evaluated and chosen in a greedy manner. The cost function that is minimized to choose split points, for regression predictive modelling problems, as in our case, is the sum squared error across all training samples that fall within the rectangle:

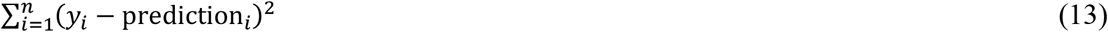

Here, *y* is the output for the training sample and prediction is the predicted output for the rectangle.

The problem with decision trees are that they are sensitive to the specific data they are trained on. Changing the training data changes the structure of the tree and hence the predictions differ. They are computationally expensive to train, with high chance of overfitting. A Random Forest is a bagging technique, where the trees are run in parallel, without any interaction. It operates by constructing a multitude of decision trees at training time and outputting mean prediction of the individual trees. A random forest is a meta-estimator which aggregates many decision trees. Here, the number of features that can be split on at each node is limited. It ensures that the ensemble model doesn’t rely too much on any individual feature. Each tree draws a random sample from the original dataset which prevents over fitting.

Ensemble Learning models have been used to test the importance of various generated features. The generated data has been used to train a Random Forest Estimator, which gave satisfactory test accuracy. Training data has been prepared by combining all the data from various states till 01/05/20and testing data included data from 02/05/20 till 09/05/20.Importance of the various features have been analysed using this Random Forest Estimator model[24]. Random forests have been used because they produce better results by ensemble the relatively uncorrelated decision trees and avoiding over-fitting [25].

Every decision tree of the forest chooses a subset of the feature set and produces a result accordingly. By analysing the result of every such decoupled decision tree, it can be decided which feature has more ability to guide us to the required result. The more a feature contributes towards minimizing the error, the more it’s importance. The minimization effect for a feature can be calculated by taking mean of the error values from the trees in which it appears. It can also be said that features that appear on higher levels of the tree are more important as they contribute to relatively higher information gain.

It can be said that random forests provide better results for feature importance as it includes results of several decision trees and thus is more generalised than other approaches.

### Steps followed

1. Data collection from Kaggle and world weather online
2. Cleaning the data and preparing a time-series dataset as suitable for our model
3. Generating new features on the dataset to analyse the relation
4. Analyse the correlation between features using Pearson Correlation Coefficient
5. Prepare a train-test dataset (Train-30/1/20-01/05/20, Test-02/05/20-09/05/20)
6. Train a random forest regression model on the training set and check for errors using the test set.
7. Extract and Analyse the Scaled Importance value of features using the trained model
8. Apply Kalman filter on the training set for future forecast of next 7 days
9. Evaluate the forecast using Mean Absolute Error

### Data Collection

Study of the COVID-19 has been done through available data in open domain. The data for number of confirmed cases of COVID-19 for India and different Indian states was collected from Kaggle website [20]. The website provides the data of covid-19 for every country [21]. The positive cases of COVID-19 in India are also collected from same [20]. The experimental data for different Indian states of COVID-19 has been taken from website [21]. It provides the different attributes like, Province/State, Country/Region, Confirmed COVID_19 cases, Death, and Cured. We collected the data between 31-01-2020 to 22-04-2020 for above study. The data of weathers are also collected for study of their effects. The maximum temperature, minimum temperature, humidity etc was fetched from available API in python. Pandas and Matplotlib in Python is used to plot these statistics.

### Data Processing

Kalman future forecast algorithm was used to predict the future growth of number of cases in India as whole, as well as Indian state wise. Kalman algorithm requires Time-Series Data as input hence some data pre-processing was done on the collected data. Pandas in Python are used to pre-process these .csv files.

As per record, the first case of Corona in India was reported on 30/01/2020 in Indian state Kerala. The Corona virus affects other states of India after few days of 1^st^ case. The present manuscript 30/1/2020 is chosen as the starting date and 22/04/2020 as the ending date for data consistency. New row data with default value for confirmed cases as zero was generated for states which reported their first case after few days. From the available features in the data, only Date and Confirmed Cases were selected, and setting the State name as index, increasing dates as columns; a time series data frame is generated which is ready to be used as input in the Kalman Algorithm. Same process was repeated for Death and Cured Cases. The statistical relationship between some of self generated features and number of confirmed cases is established with the help of ensemble-learning models. The features generated to analysis of effect of different parameters in present manuscripts are Confirmed Cases 1 day ago, Growth in 1 day, Growth rate in 1 day (100 ∗ (*Growthin*1*day*) ÷ (*Cases*1*dayago*)), Growth in 3 days, Growth rate in 3 days, Growth in 5 days, Growth rate in 5 days, Growth in 7 days, Growth rate in 7 days, Max Temperature in C, Min Temperature in C and Humidity. Table 1 represents the temperature and humidity data, during the study period, of 15 states of India chosen for this study. Table 2 represents different features of the data collected for spread scenario.

**Table 1:** Temperature and Humidity data of 15 States of India.

| S. No | State | Average min Temp | Average max Temp | Average Humidity |
| --- | --- | --- | --- | --- |
| 01 | Andhra Pradesh | 27 | 31 | 73 |
| 02 | Delhi | 28 | 37 | 24 |
| 03 | Gujarat | 28 | 41 | 31 |
| 04 | Haryana | 20 | 33 | 32 |
| 05 | Jammu & Kashmir | 7 | 19 | 62 |
| 06 | Karnataka | 24 | 35 | 33 |
| 07 | Madhya Pradesh | 25 | 39 | 21 |
| 08 | Maharashtra | 30 | 34 | 57 |
| 09 | Punjab | 20 | 33 | 32 |
| 10 | Rajasthan | 24 | 35 | 29 |
| 11 | Tamil Nadu | 26 | 32 | 69 |
| 12 | Telangana | 29 | 40 | 27 |
| 13 | Uttar Pradesh | 27 | 37 | 24 |
| 14 | West Bengal | 14 | 37 | 66 |
| 15 | Kerala | 35 | 27 | 71 |

**Table 2:** COVID-19 data of 15 states of India.

| Date<br>(2020) | State | Confirmed<br>Cases | Cases<br>1 day<br>ago | Growth<br>in 1<br>day | Growth<br>in 3<br>days | Growth<br>in 5<br>days | Growth<br>in 7<br>days | Growth<br>rate for<br>1 day | Growth<br>rate for<br>3 days | Growth<br>rate for<br>5 days | Growth<br>rate for<br>7 days |
| --- | --- | --- | --- | --- | --- | --- | --- | --- | --- | --- | --- |
| 04-21 | Andhra Pradesh | 757 | 722 | 35 | 154 | 223 | 284 | 4.84 | 25.53 | 41.76 | 60.04 |
| 04-21 | Delhi | 2081 | 2003 | 78 | 374 | 503 | 571 | 3.89 | 21.90 | 31.87 | 37.81 |
| 04-21 | Gujarat | 2066 | 1851 | 215 | 794 | 1195 | 1449 | 11.61 | 62.42 | 137.19 | 234.84 |
| 04-21 | Haryana | 254 | 233 | 21 | 29 | 49 | 55 | 9.01 | 12.88 | 23.90 | 27.63 |
| 04-21 | Jammu & Kashmir | 368 | 350 | 18 | 40 | 68 | 98 | 5.14 | 12.19 | 22.66 | 36.29 |
| 04-21 | Karnataka | 415 | 395 | 20 | 44 | 100 | 157 | 5.06 | 11.85 | 31.74 | 60.85 |
| 04-21 | Kerala | 408 | 402 | 6 | 12 | 20 | 29 | 1.49 | 3.03 | 5.15 | 7.65 |
| 04-21 | Madhya Pradesh | 1540 | 1485 | 55 | 185 | 420 | 810 | 3.70 | 13.65 | 37.5 | 110.95 |
| 04-21 | Maharashtra | 4669 | 4203 | 466 | 1346 | 1750 | 2332 | 11.08 | 40.50 | 59.95 | 99.78 |
| 04-21 | Punjab | 245 | 219 | 26 | 43 | 59 | 69 | 11.87 | 21.28 | 31.72 | 39.20 |
| 04-21 | Rajasthan | 1576 | 1478 | 98 | 347 | 553 | 697 | 6.63 | 28.23 | 54.05 | 79.29 |
| 04-21 | Tamil Nadu | 1520 | 1477 | 43 | 197 | 278 | 347 | 2.91 | 14.89 | 22.384 | 29.58 |
| 04-21 | Telengana | 919 | 873 | 46 | 128 | 221 | 295 | 5.26 | 16.18 | 31.661 | 47.275 |
| 04-21 | Uttar Pradesh | 1294 | 1176 | 118 | 325 | 521 | 637 | 10.03 | 33.53 | 67.39 | 96.95 |
| 04-21 | West Bengal | 392 | 339 | 53 | 105 | 161 | 202 | 15.63 | 36.58 | 69.69 | 106.31 |

The authors for this manuscript collected the data of the temperature and humidity from fetched https://www.worldweatheronline.com/ API in python. All the major COVID-19 hotspots cities were considered from each states and an average of their data was allotted to the corresponding state.

### Results and discussion

The proposed methodology has been validated with the cases reported over 15 different states of India. These states of India have been chosen for this study due to large number of COVID-19 cases has been reported there. We have chosen 15 top states according to number of cases reported. These states are Andra Pradesh, Delhi, Kerala, Madhya Pradesh, Jammu and Kashmir, Haryana, Karnataka, Gujarat, Maharashtra, Punjab, Rajasthan, Telengana, Tamil Nadu, Uttar Pradesh and West Bengal. We have taken number of total positive cases of Covid-19 from all these states for our analysis as well as prediction purpose. Training of the proposed model has been done by using the data between January 30 and May 09, 2020. Figure 1 shows the spread scenario in these states during this period.

**Figure 1:**
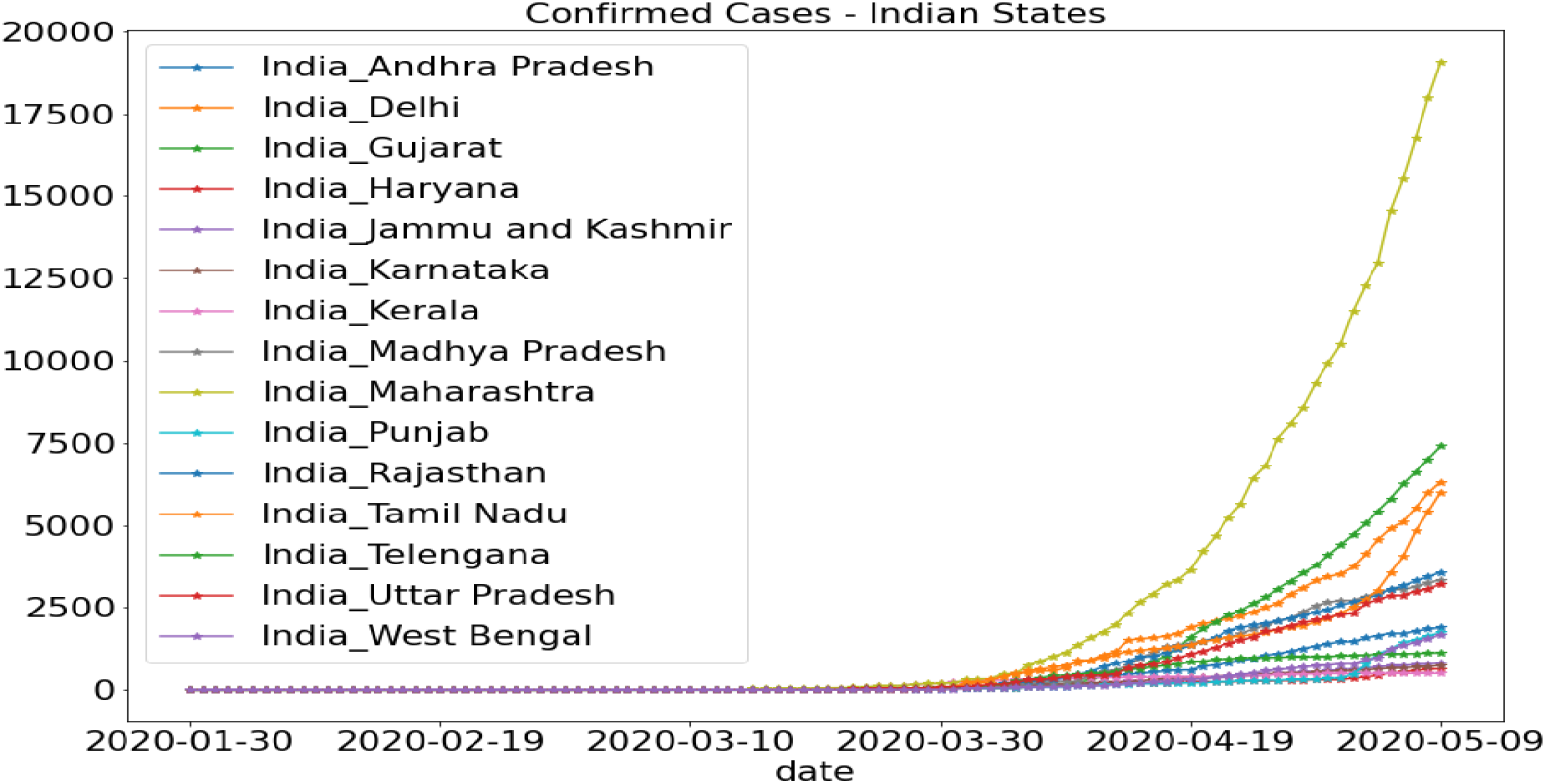
COVID-19 Spread Scenario if different Indian States

After cleaning and preparing the dataset, *Pearson correlation* has been calculated for each pair of features using equation 12 and the results have been analysed using *Heat map*. Each column of the heat map is representing the dependency of the X-axis parameter on the Y-axis parameters. Heat map of the Pearson Coefficient for pair of features for COVID-19 cases has been shown in fig. 2. It has been observed from fig 2 that confirmed cases have strong positive correlation with growth in 1 day, growth in 3 days, growth in 5 days, and growth in 7 days. Total confirmed cases are highly dependent on the cases reported in previous 7 days. Hence, it is supporting the well exist idea of COVID-19 spread chain which is dependent on number of previous cases [22, 23].

**Figure 2:**
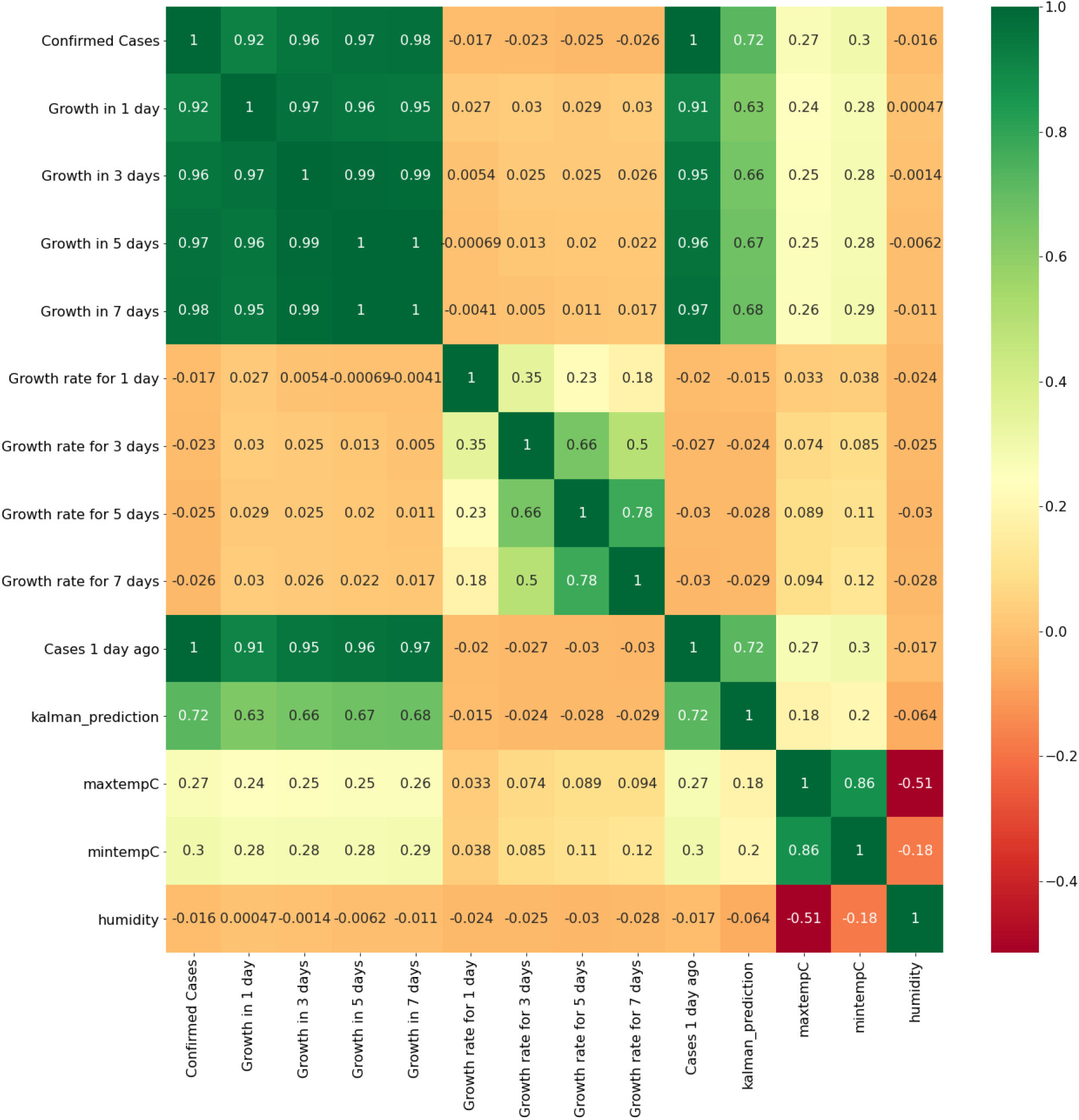
Heat Map of Pearson Coefficients for COVID-19 Spread

It has been also noted that prediction model is also having positive correlation with previous day cases. The effect of historical data about the spread has less correlation in prediction as compared to previous day data. It can be seen from fig 2 that minimum temperature and maximum temperature are having weak positive correlation in the spread. It has been seen from heat map that minimum temperature is more crucial than maximum temperature in spread analysis.

Train-test split is done on dataset based on date. Training data belongs to data from January 30, 2020 to May 01, 2020. Validation data has been chosen from May 02–09, 2020. The input features are Confirmed Cases 1 day ago, Growth in 1 day, Growth in 3 days, Growth in 5 days, Growth in 7 days, Growth rate in 1 day, Growth rate in 3 days, Growth rate in 5 days, Growth rate in 7 days, Maximum Temperature in Centigrade, Minimum Temperature in Centigrade and Humidity and the output is Confirmed Cases. This dataset is then fitted into the Random Forest Regression Model. On evaluating, test dataset gives a Mean Absolute Error of 109.85. This model is then used to analyse the feature importance of different features on the target variable.

Figure 3 represents the importance of different features on total confirmed cases by using random forest method. It has been noted from fig 3 that historical spread data has high importance for the prediction. It has also been seen from fig 3 that maximum temperature is having very less importance as compared to minimum temperature. Humidity is also playing crucial role in spread and is well noted from the figure. It has been pointed out from fig. 2 and 3 that humidity is having negative correlation with confirmed cases and prediction model also. It has higher importance for prediction than temperature.

**Figure 3:**
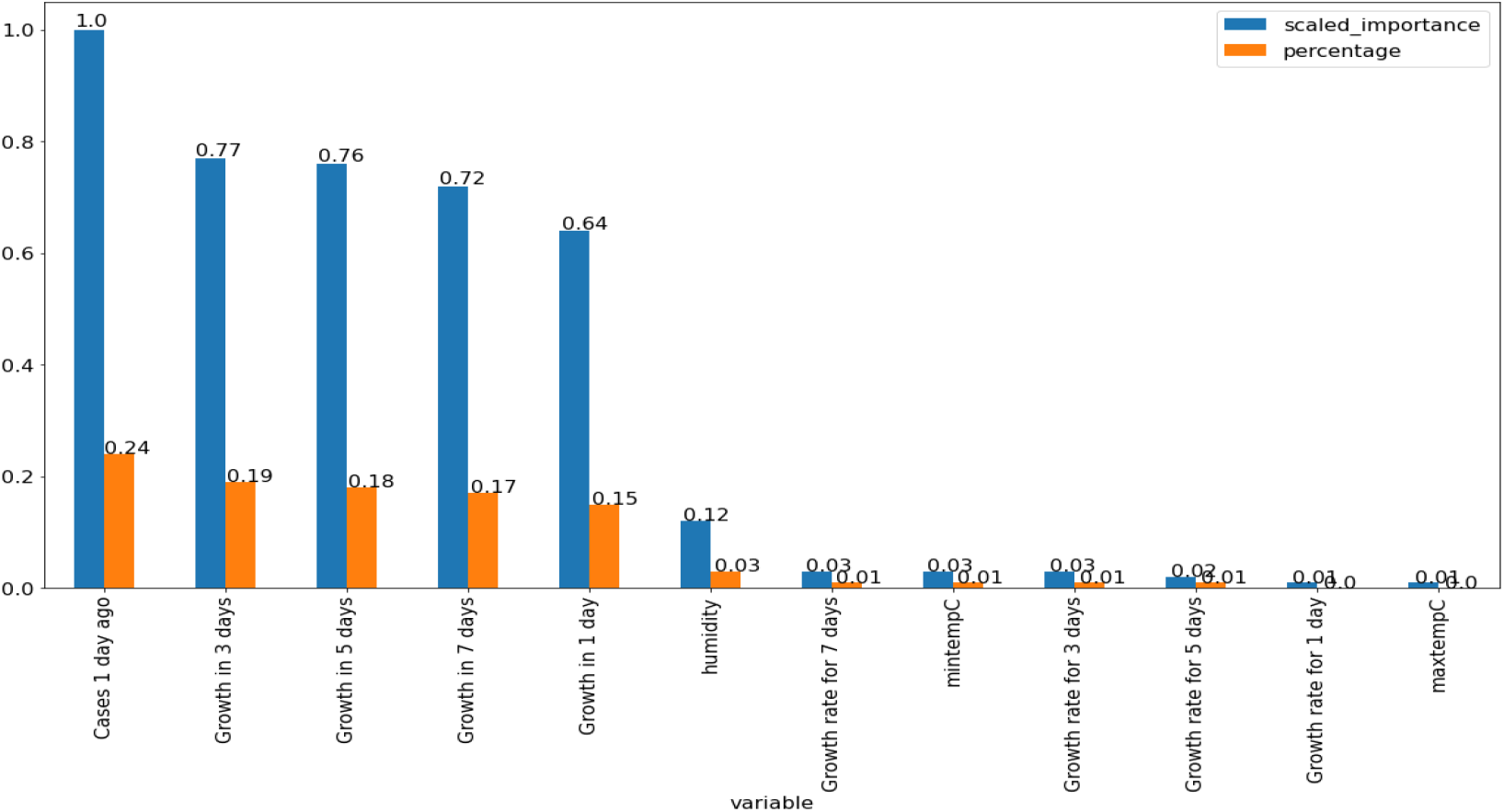
Scaled Importance of features in COVID-19 spread using Random Forest

The time-series dataset prepared is then used for future forecast of COVID-19 cases; state-wise first then for India. Firstly, our Kalman Filter is applied for the training till May 01, 2020. Validation of our prediction model has been performed by using the dataset fro May 02–09, 2020. Predicted data has been compared with the real data for same time period. Table 3 shows the average mean error reported for different states of India in prediction. This error is absolute difference between predicted values and real data. It can be noted that mean average error is varying in the range of 24 to 1297 for different states. It has been pointed out from table 3 that the validation results are very good except few states like Tamilnadu, Panjab, Maharastra and Gujrat. These states are showing different behaviour of COVID-19 spread from the prediction. The reason behind this deviation is the delay in declaration of testing results. Mass results have been declared in one day. Hence, they are showing different behaviour.

**Table 3:** Mean Average Error in Validation of prediction model state wise.

| S. No | State | MAE(1day) | MAE(7day) | MAE(15days) |
| --- | --- | --- | --- | --- |
| 01 | Andhra Pradesh | 12.00 | 46.42 | 55.336 |
| 02 | Delhi | 7.00 | 42.42 | 1599.27 |
| 03 | Gujarat | 35.00 | 137.42 | 195.46 |
| 04 | Haryana | 24.00 | 151.00 | 68.27 |
| 05 | Jammu &Kashmir | 1.00 | 24.71 | 131.6 |
| 06 | Karnataka | 25.00 | 45.71 | 38.8 |
| 07 | Madhya Pradesh | 26.00 | 141.42 | 162.2 |
| 08 | Maharashtra | 128.00 | 337.14 | 3527.06 |
| 09 | Punjab | 133.00 | 814.14 | 440.80 |
| 10 | Rajasthan | 29.00 | 46.85 | 340.93 |
| 11 | Tamil Nadu | 372.00 | 1297.71 | 1214.20 |
| 12 | Telengana | 1.00 | 52.57 | 145.27 |
| 13 | Uttar Pradesh | 7.00 | 38.57 | 245.40 |
| 14 | West Bengal | 16.00 | 82.71 | 375.20 |
| 15 | Kerala | 2.00 | 35.57 | 38.80 |

Figure 4 shows the validation results of prediction model with state wise data. Some random states have been chosen to show the graphical results. All state data has been shown in table 3.

**Figure 4:**
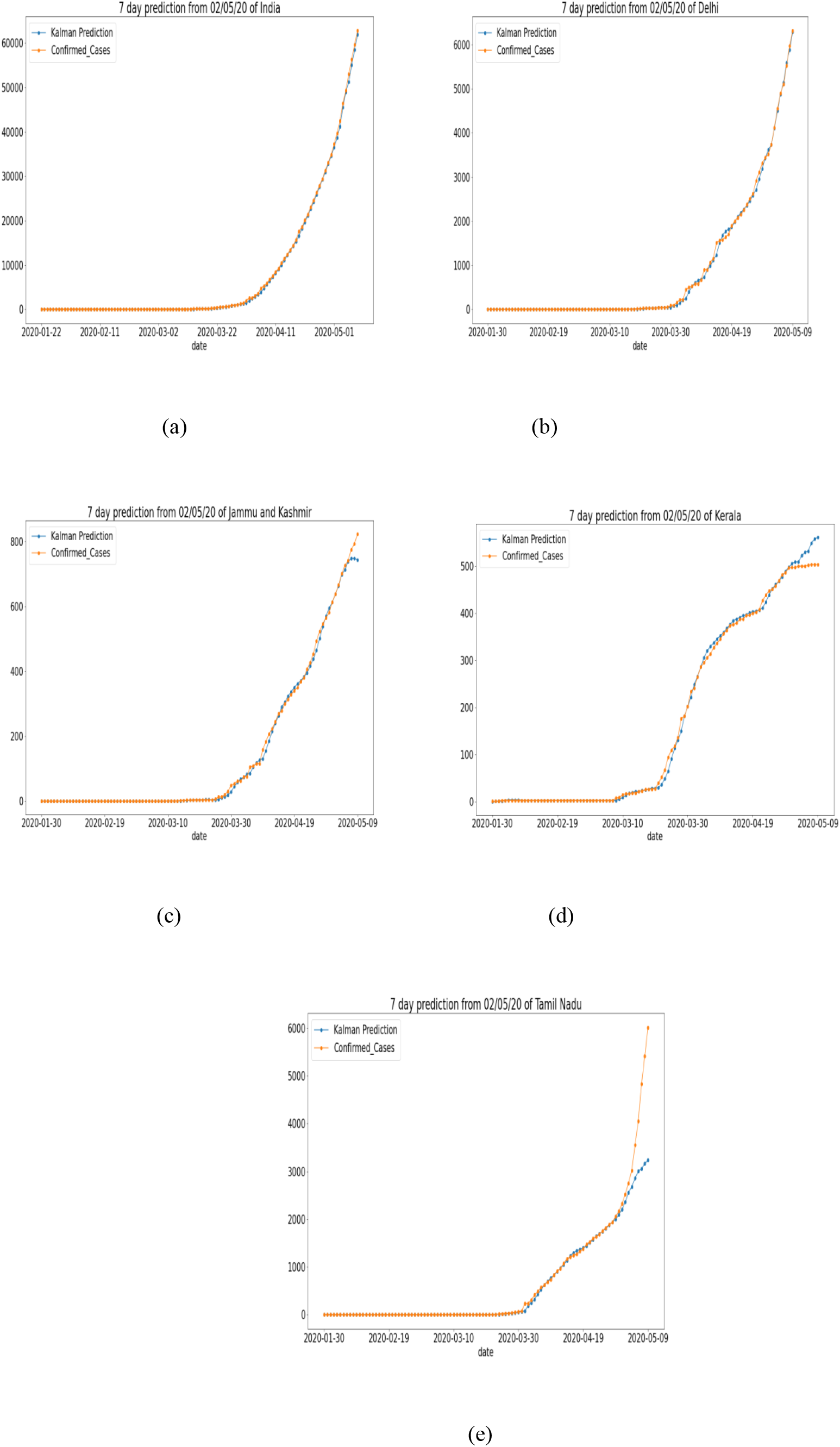
Validation Results of Prediction model for different states

Figure 5 shows the results obtained by prediction model for next 30 days cases of COVID-19 for different states in India. It has been observed that Kalman filter based prediction model shows higher deviation from real data for long term prediction. Kalman filter based prediction is more accurate for short term prediction. This phenomenon is well supported by Pearson coefficient and random forest based study. Both the studies are showing that confirmed cases have strong positive correlation as well as high importance for historical spread data. Hence, any error in prediction for a single day will be propagated and will produce the larger error after few days. Hence, Kalman filter based prediction model is good for short term prediction i.e. Daily and Weekly.

**Figure 5:**
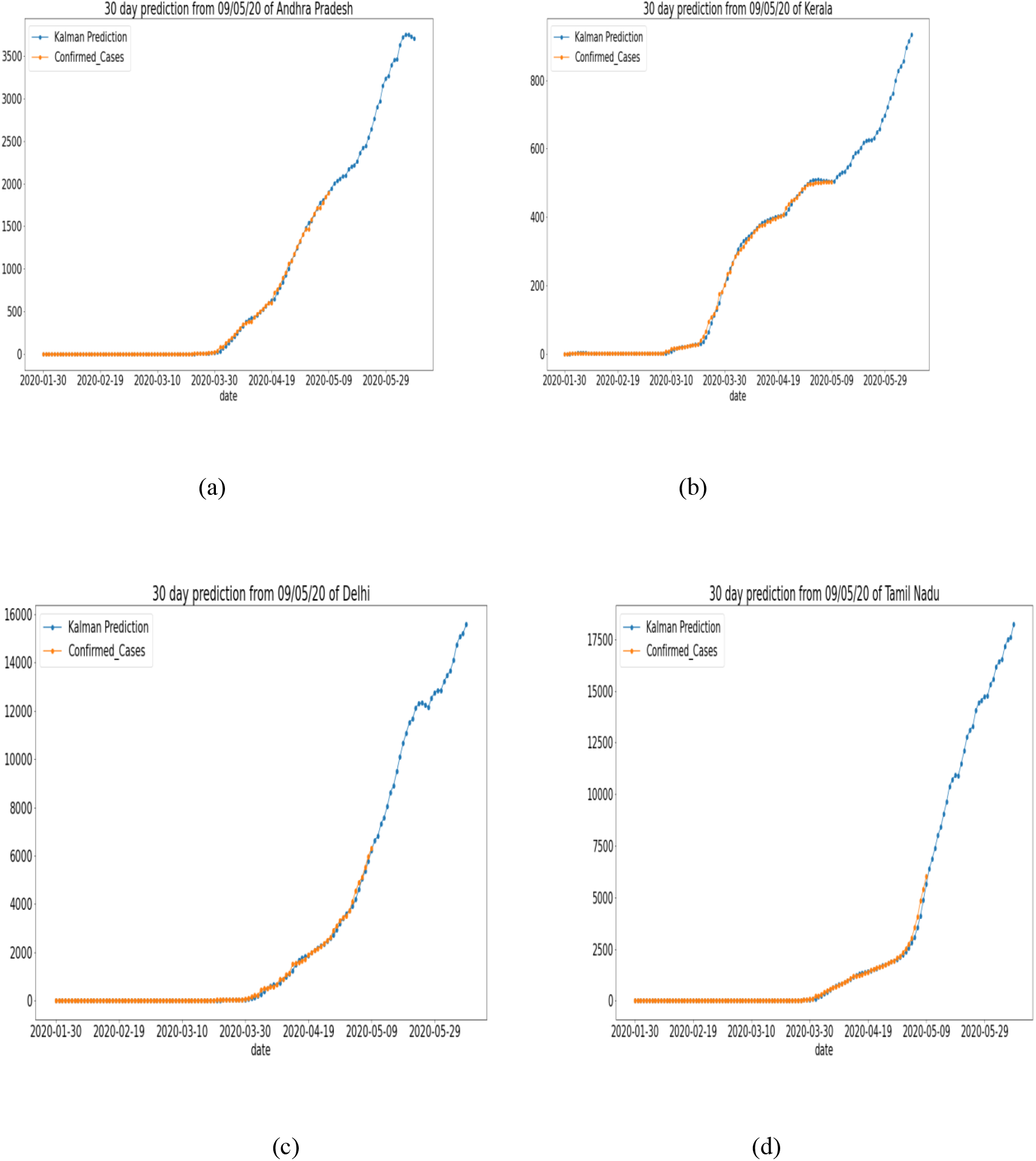
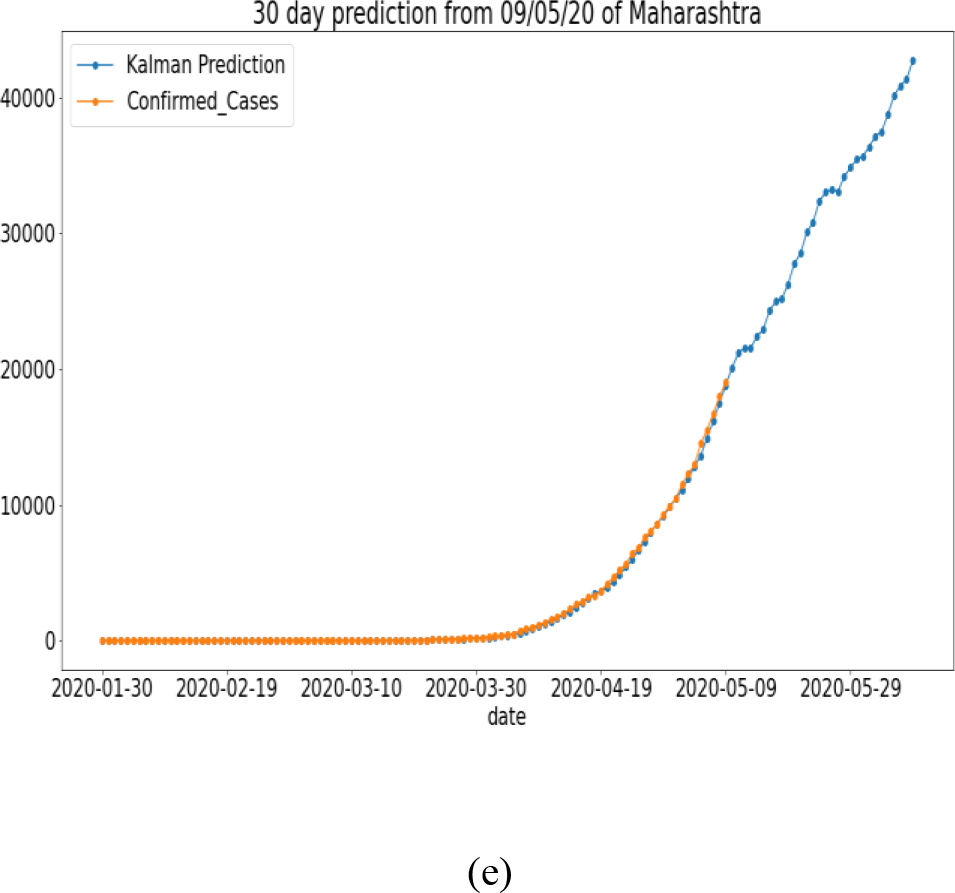
prediction results for next 30 days for different states of India

## Conclusions

The present manuscript presented a prediction model based on Kalman filter. The correlations between different features of COVID-19 spread have been studied. It has been found that previous spread data has strong positive correlation with the prediction. The importance of different features in prediction model has also been studied in the present manuscript. It has been noted that historical spread scenario has large impact on the current spread. Hence, it can be concluded that COVID-19 spread is following a chain. Hence, to reduce the spread this chain has to be breaked. The proposed prediction model is providing encouraging results for the short term prediction. It has been noted that for long term prediction, Kalman filter based proposed model is showing large mean average error. Hence, it can be conclude that proposed prediction model is good for short term prediction i.e. daily and weekly. The proposed prediction model can be updated to accommodate long term and medium term time series prediction in future.

## Data Availability

Code will be available on request

